# Influence of temperature and precipitation on the effectiveness of water, sanitation, and handwashing interventions against childhood diarrheal disease in rural Bangladesh: a re-analysis of a randomized control trial

**DOI:** 10.1101/2022.09.25.22280229

**Authors:** Anna T. Nguyen, Jessica A. Grembi, Marie Riviere, Gabriella Barratt Heitmann, William D. Hutson, Tejas S. Athni, Arusha Patil, Ayse Ercumen, Audrie Lin, Yoshika Crider, Andrew Mertens, Leanne Unicomb, Mahbubur Rahman, Stephen P. Luby, Benjamin F. Arnold, Jade Benjamin-Chung

**Author notes:** Corresponding Author: Anna T. Nguyen. 300 Pasteur Drive, Palo Alto, California 94305. Conflicts of Interest Statement: The authors declare they have nothing to disclose.

## Abstract

**Background:** Diarrheal disease is a leading cause of childhood morbidity and mortality globally. Household water, sanitation, and handwashing (WASH) interventions can reduce exposure to diarrhea-causing pathogens, but climatic factors may impact their effectiveness. Information about effect heterogeneity under different weather conditions is critical to intervention targeting.

**Methods:** We analyzed data from a trial in rural Bangladesh that compared child diarrhea prevalence between clusters that were randomized to different WASH interventions between 2012-2016 (NCT01590095). We matched temperature and precipitation measurements to households by geographic coordinates and date. We estimated prevalence ratios (PR) using generative additive models and targeted maximum likelihood estimation to assess the effectiveness of each WASH intervention under different environmental conditions.

**Findings:** Generally, WASH interventions most effectively prevented diarrhea during monsoon season, particularly following weeks with heavy rain or high temperatures. Compared to the control arm, WASH interventions reduced diarrhea by 51% (95% CI 33%-64%) following periods with heavy rainfall vs. 13% (95% CI -26%-40%) following periods without heavy rainfall. Similarly, WASH interventions reduced diarrhea by 40% (95% CI 16%-57%) following above-median temperatures vs. 17% (95% CI -38%-50%) following below-median temperatures. The influence of precipitation and temperature varied by intervention type; for precipitation, the largest differences in effectiveness were for the sanitation and combined WASH interventions.

**Interpretation:** WASH intervention effectiveness was strongly influenced by precipitation and temperature, and nearly all protective effects were observed during the rainy season. Future implementation of these interventions should consider local environmental conditions to maximize effectiveness.

**Funding:** Bill & Melinda Gates Foundation; National Institute of Allergy and Infectious Diseases; National Heart, Lung, And Blood Institute; National Institute of General Medical Sciences; Stanford University School of Medicine; Chan Zuckerberg Biohub

**Research in Context Panel:** *Evidence before this study:* We searched Google Scholar using the search terms “sanitation” OR “hygiene” OR “WASH” OR “water quality”; AND “heterogen*” OR “effect modif*”; AND “temperature” OR “precipitation” OR “rain*” OR “climate” OR “environmental”; AND “diarrhea” OR “enteric infection”; AND “risk” AND/OR “factors”. In general, the effect modification of WASH interventions on diarrhea by weather is not well studied. One study in Ecuador investigated different relationships between rainfall, diarrhea, and unimproved sanitation and water sources. They found that unimproved sanitation was most strongly associated with elevated diarrhea after low rainfall, whereas unimproved water sources were most strongly associated with elevated diarrhea after heavy rainfall. In a similar setting in Ecuador, a separate study found that drinking water treatments reduced increases in diarrhea after heavy rainfall that followed dry periods, while sanitation and hygiene had no impact on the relationship between heavy rainfall and diarrhea. One study in Rwanda also found that high levels of runoff were protective against diarrhea only in households with unimproved toilets. In Bangladesh, one study found that access to tubewells was most effective at reducing childhood diarrhea in non-flood controlled areas. High heat can accelerate the inactivation of enteric pathogens by water chlorination, but no studies have examined how temperature influences the effectiveness of sanitation or hygiene interventions. No prior studies have estimated differences in WASH effectiveness under varying weather conditions within a randomized trial.

*Added value of this study:* To our knowledge, this is the first study to assess differences in household-level WASH intervention effectiveness by weather conditions in a randomized trial. We spatiotemporally matched individual-level data from a trial in rural Bangladesh to remote sensing data on temperature and precipitation and estimated differences in the effectiveness of WASH interventions to prevent childhood diarrhea under varying levels of these environmental factors.

*Implications of all the available evidence:* We found that WASH interventions were substantially more effective following periods with higher precipitation or higher temperatures. We observed the largest effect modification by precipitation for a sanitation intervention. This may be because compared to water and handwashing interventions, the sanitation intervention blocked more pathways through which enteric pathogens reach water, soil, and flies following heavy rainfall. In regions like Bangladesh, extreme weather is expected to become more common under climate change but WASH interventions might mitigate increases in childhood diarrhea due to climate change.

## Background

In 2019, over 500,000 under-5 child deaths were caused by diarrheal disease.^1^ Children that suffer from repeated diarrheal episodes are at high risk of malnutrition, stunted growth, and impaired cognitive development.^2^ The World Health Organization estimates that over half of diarrhea deaths are directly attributable to inadequate water safety, sanitation, and/or handwashing (WASH).^3^ Low-cost, household level WASH interventions may prevent the spread diarrhea-causing pathogens, leading to improvements in child growth and development.^2^ However, randomized controlled trials of WASH interventions in rural Kenya and Bangladesh found surprisingly modest effects on diarrhea; in Bangladesh, there was a 39% reduction in diarrhea prevalence among children who received a combined water, sanitation, and hygiene intervention, and in Kenya there was no reduction.^4, 5^

Diarrheal disease is associated with temperature^6–10^ and precipitation^7–11^ across many different settings, and there are multiple pathways through which environmental conditions might influence the relationship between WASH interventions and diarrhea. Each type of WASH intervention prevents different subsets of enteric pathogen transmission pathways, and each may be distinctly influenced by weather.^12^ Heavy rainfall may result in damage to latrines, causing feces to contaminate the household environment and nearby food and water sources.^13^ During heavy rainfall events, it may be more difficult for parents to use child potties and to scoop child feces and place them in latrines. Under higher temperatures, the survival of certain pathogens on surfaces and in water sources may be higher^14^, and low-cost handwashing interventions and water chlorination and safe storage may be insufficient to prevent diarrhea.

Yet, few studies have examined how climate and environmental factors modify the effect of WASH interventions on diarrhea. There is some evidence that the relationships between diarrhea prevalence and unimproved water sources and sanitation systems changed under different levels of precipitation and runoff ^11, 15^, but that water treatment could mitigate increases in diarrhea following heavy rainfall in some conditions.^16^ In Bangladesh, one study found that tubewells were most effective in non-flood controlled areas.^17^ Another study found that reductions in diarrhea from sanitation interventions occurred exclusively during the rainy season in Bangladesh.^18^ However, most prior studies used observational designs, and estimates of WASH intervention effectiveness were likely to be confounded by household wealth.

Our objective was to assess whether temperature and precipitation modified the effect of water, sanitation, and handwashing interventions on child diarrhea prevalence in a randomized trial in rural Bangladesh. We merged individual-level outcome data with granular weather measurements from remote sensors to model how environmental conditions influence intervention effectiveness. Understanding the impact of weather on intervention effects may help guide the targeted implementation of future WASH interventions.

## Methods

### Study Data

The WASH Benefits Bangladesh trial (NCT01590095) delivered low-cost, household-level WASH interventions and estimated their effects on diarrheal disease.^5, 19^ The trial enrolled pregnant women in their second or third trimester between 2012-2013 in the Gazipur, Kishoreganj, Mymensingh, and Tangail districts of rural Bangladesh. Village clusters were block-randomized to one of the following arms: chlorinated drinking water (W); upgraded sanitation (S); promotion of handwashing with soap (H); nutrition education with lipid-based supplement (Nutrition); combined water, sanitation, and handwashing (WSH); combined water, sanitation, handwashing, and nutrition (WSH + Nutrition); or control. In total, there were 720 village clusters included in the trial; 90 clusters were randomized to each intervention arm and 180 clusters were randomized to a double-sized control arm. Here, we excluded the Nutrition arm from this analysis since the effectiveness of nutrition alone is unlikely to depend on environmental factors. For similar reasons, we merged the WSH and WSH + Nutrition groups to a single “combined WASH” group.

The water intervention included a lidded storage container and regular supply of sodium dichloroisocyanurate tablets. The sanitation intervention included the installation of a double-pit pour-flush latrine compared to the more common single-pit pour-flush latrine. Fecal matter can be diverted to a secondary chamber for compost in the double-pit latrine, while waste needs to be manually removed from the single-pit latrine. The handwashing intervention included numerous handwashing stations and a regular supply of detergent sachets to make soapy water. All interventions were delivered by community promoters who provided instruction for proper use.

There were 4,747 children born to the enrolled mothers (“index children”). In the study region, children lived within compounds shared by their extended family that consist of their own household (the “index household”) and an average of 1.5 other households (range 0-10). The water and handwashing interventions were only implemented in the index household, while the sanitation intervention was provided to all households in the compound.

Additional details on enrollment criteria and intervention details have been reported elsewhere.^5^

### Outcome Data

Diarrhea was measured for index children and children living in the same compound who were under the age of three at enrollment. Measurements were taken approximately 1 and 2 years after intervention delivery through caregiver report of at least three loose stools in 24-hours or a bloody stool within the past 7 days.

### Environmental Data

We assessed pre-specified measures of precipitation and temperature as possible effect modifiers. We matched precipitation and temperature values from spatiotemporal remote sensing data to each trial measurement by household coordinates and the date of outcome assessment. We calculated distances between sensor measurements and study coordinates using the haversine formula.

#### Precipitation

We obtained precipitation data from the Multi-Source Weighted-Ensemble Precipitation dataset from GloH20 (daily temporal resolution, 0.1° spatial resolution), which merges numerous gauge, satellite, and reanalysis precipitation data sources and corrects for bias.^20^ We calculated the total weekly precipitation, and created binary indicators of whether these sums exceeded the median weekly total precipitation across the study period. To measure heavy rain, we created binary indicators for at least one day in the week in which total precipitation was above the 80^th^ percentile of all daily totals.

We defined the rainy season as the continuous period during which the 5-day rolling average of daily precipitation was over 10 mm, and constructed variables to indicate if diarrhea measurements were taken during the rainy season.

#### Temperature

We obtained daily near surface air temperature data from the Famine Early Warning Systems Network Land Data Assimilation System (FLDAS) Central Asia dataset (daily temporal resolution, 0.01° spatial resolution) from the National Aeronautics and Space Administration (NASA).^21^ We computed the minimum, maximum, and average temperatures. We also constructed binary indicators for whether the minimum, maximum, and average temperatures exceeded the median value across the study period.

#### Lag Periods

Our primary analysis used weekly measures of temperature and precipitation with a 1-week lag (capturing the 8-14 day period prior to date of caregiver reported illness) to account for the period of time in which weather conditions could influence enteric pathogen transmission in the environment and the short incubation period of common enteric pathogens.^22^ We conducted sensitivity analyses using alternative lag periods: 0 weeks (1-7 days prior), 2 weeks (8-21 days prior), and 3 weeks (22-28 days prior).

### Statistical Analysis

Intervention adherence was high in the trial^5^, so we conducted an intention-to-treat-analysis, we estimated the effect of WASH interventions on diarrhea prevalence under different environmental conditions. We compared diarrhea prevalence in the control group to those receiving the following interventions: (1) water only, (2) sanitation only, (3) handwashing only, (4) combined WASH (WSH or WSH + Nutrition), and (5) any WASH intervention (water, sanitation, hygiene, combined WSH, or combined WSH + Nutrition arms).

To allow for potential non-linear relationships between continuous weather measures and intervention effects, we fit generalized additive models (GAMs) to model the outcomes as a function of the intervention and an environmental variable (as a spline). We specified a binomial family with logit link functions and used restricted maximum likelihood estimation to select smoothing parameters. We estimated simultaneous confidence intervals using a parametric bootstrap of the variance-covariance matrix under a multivariate normal distribution. Additionally, we estimated prevalence ratios for any intervention vs. control at the 10^th^ and 90^th^ percentile of the observed environmental variables and calculated the corresponding 95% confidence intervals using a non-parametric bootstrap with resampling at the cluster level. We estimated intervention effectiveness under categorical weather measures using targeted maximum likeli estimated the prevalence ratios and corresponding confidence intervals for intervention vs. control for each weather variable stratum. We used cluster-level influence curve-based standard errors to account for dependence within village clusters.

We adjusted the models to consider the relationships between temperature and precipitation. In models where temperature was the effect modifier, we controlled for total weekly precipitation. In models where precipitation was the effect modifier, we controlled for average weekly temperature.

To check for possible misclassification of reported diarrhea, we conducted a negative control analysis using caregiver-reported bruising in the past 7 days.

Analyses were conducted in R (version 4.1.3). The pre-analysis plan and replication scripts are available through the Open Science Framework (https://osf.io/yt67k/). We note deviations from the pre-analysis plan in Appendix 1.

### Role of the Funding Source

The funders of the study had no role in study design, data analysis, interpretation, writing of the manuscript, or the decision to submit the manuscript for publication.

## Results

Our analysis included 12,440 total diarrhea measurements for 6,921 children between 0.5 and 5.5 years of age (mean 2.0, SD 1.2) during the period between February 03, 2013 and October 31, 2015. (Figure 1) We report additional participant characteristics and the distribution of environmental factors by group in Table 1.

**Figure 1:**
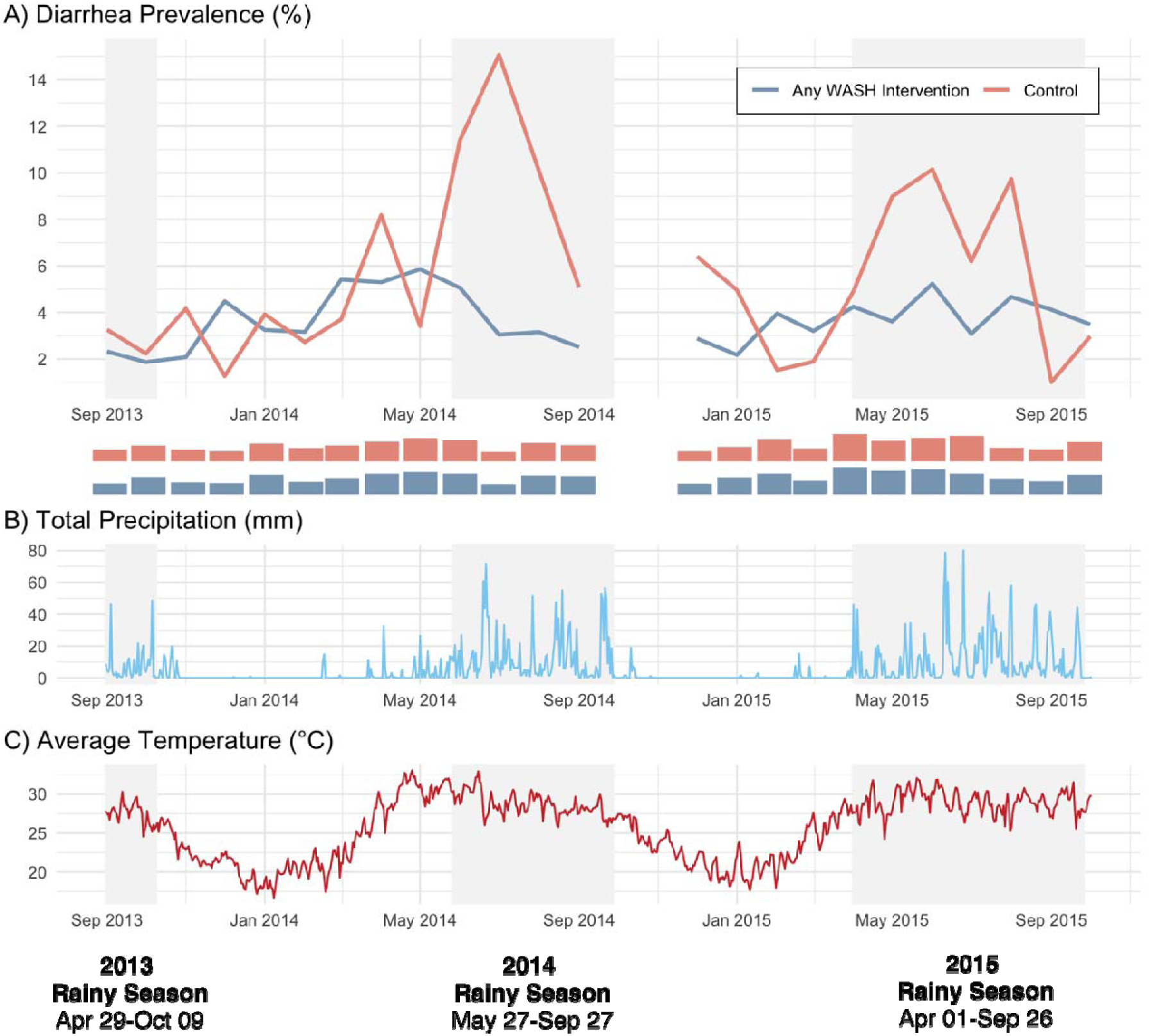
Diarrhea prevalence, precipitation, and temperature over time. A) Prevalence of caregiver-reported diarrhea over time, by intervention group. Rainy season is shaded in gray. Rug plots show the number of diarrhea measurements made per month during the trial. Plot is left-truncated at September 2013, such that the figure omit two measurements taken in March 2013 that are included in subsequent analyses. B) Daily total precipitation over time, averaged across the study area C) Daily average temperature over time, averaged across the study area

**Table 1:** Population characteristics by intervention group. Sample sizes, child demographics, diarrhea prevalence, and environmental risk factor distributions, by treatment arm. For categorical variables, the number of occurrences and percentages are reported. For continuous variables, the mean and 95% confidence interval are reported. All environmental variables are reported for the 8-14 day period prior to outcome assessment (1-week lag), with the exception of rainy season.

| **Variable** | **Any WASH Intervention** | **Control** |
| --- | --- | --- |
| **Sample Characteristics** |  |  |
| Children | 1,956 | 4,965 |
| Observations | 3,466 | 8,974 |
| Households | 1,203 | 3,051 |
| Children per household | 1.63 (1.57 - 1.68) | 1.63 (1.59 - 1.66) |
| Age at first measurement (years) | 1.63 (1.58 - 1.68) | 1.59 (1.55 - 1.62) |
| Age at second measurement (years) | 2.52 (2.47 - 2.58) | 2.53 (2.5 - 2.57) |
| Diarrhea Prevalence | 197 (10.1%) | 343 (6.9%) |
| **Temperature Variables** |  |  |
| Mean weekly temperature (°C) | 23.5 (23.1 - 23.9) | 23.5 (23.3 - 23.8) |
| Minimum weekly temperature (°C) | 15.8 (15.4 - 16.2) | 15.7 (15.5 - 16) |
| Maximum weekly temperature (°C) | 30.6 (30.3 - 31) | 30.6 (30.4 - 30.8) |
| **Precipitation Variables** |  |  |
| Measured during rainy season | 1772 (51.13%) | 4578 (51.01%) |
| Mean weekly total precipitation (mm) | 41.5 (32.4 - 50.7) | 41.7 (36.1 - 47.4) |
| Heavy rain (1+ days ≥ 80th percentile) | 1531 (44.17%) | 3942 (43.93%) |

### Precipitation

During the study, the total weekly precipitation ranged from 0 to 295 mm with a median of 13 mm. Precipitation was highly concentrated during the rainy season, which fell between April 29-October 9 in 2013, May 27-September 27 in 2014, and April 1-September 26 in 2015. In the control arm, we observed increases in diarrhea prevalence during the rainy seasons, particularly in 2014. In households that received any WASH intervention, diarrhea prevalence remained relatively constant over time. (Figure 1A). We saw that increases in diarrhea coincided with periods that experienced the most rainfall, with annual precipitation being highly concentrated in the rainy season. (Figure 1B)

In weeks with total precipitation over 50 mm, diarrhea prevalence was higher in the control arm than in the intervention arms, and prevalence in the control arm increased with precipitation (Figure 2). The prevalence ratio associated with any WASH intervention vs. control was 1.36 (95% CI 1.06-1.72) at the 10^th^ percentile of total rainfall (0 mm) vs 0.66 (95% CI 0.4-1.22) at the 90^th^ percentile (125 mm).

**Figure 2:**
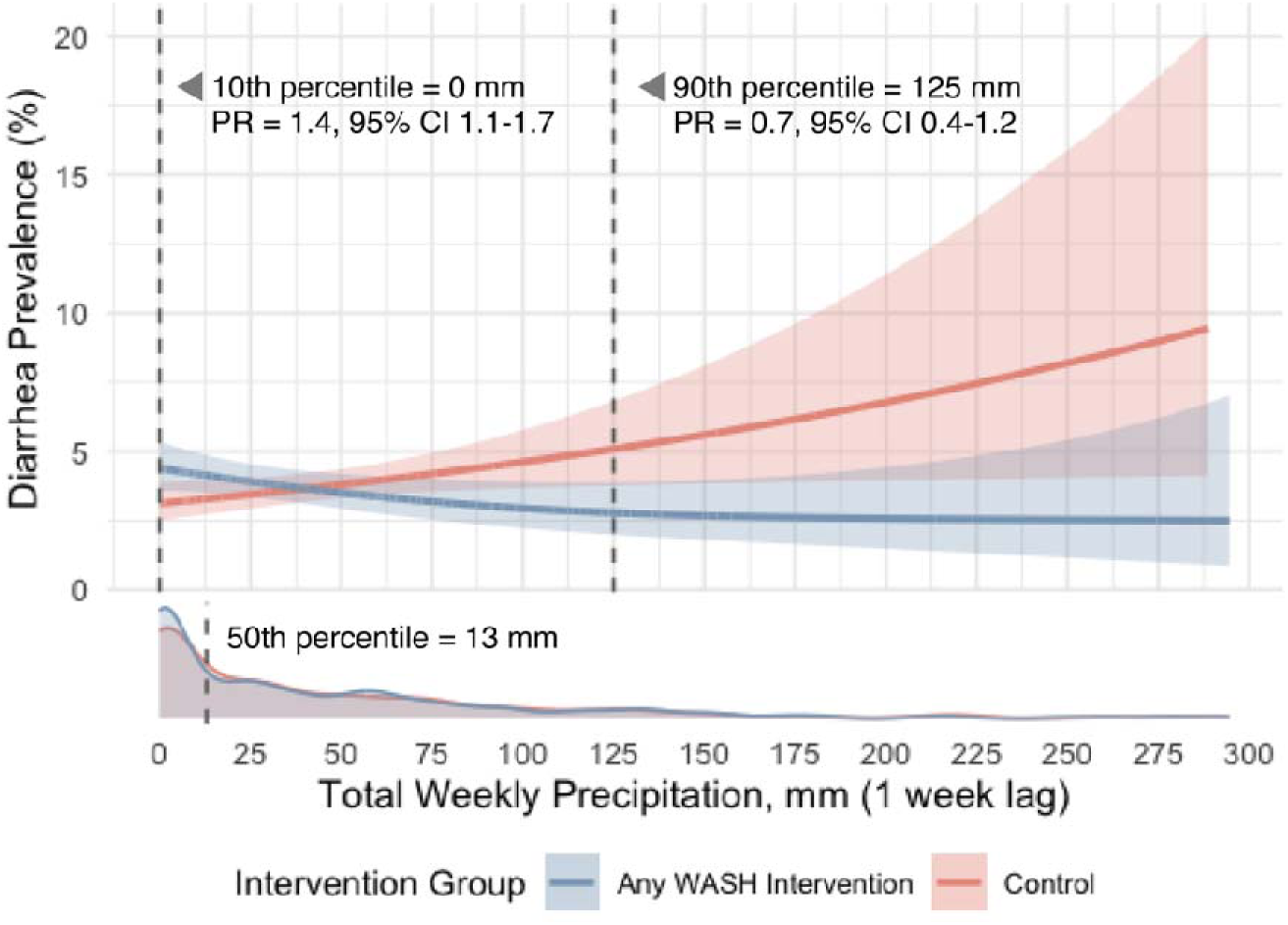
Diarrhea prevalence by total precipitation and intervention group. Predicted diarrhea prevalence by total precipitation in the 8-14 day period prior to outcome assessment, controlled for average temperature and stratified by intervention group. Prevalence ratios for the intervention are calculated at the 10th and 90th percentile of total weekly precipitation using a non-parametric bootstrap with 1,000 resamples taken at the cluster level. The density plot shows the distribution of measurements over values of weekly total precipitation, with a dashed line marking the median.

First, we examined effect modification of precipitation pooling across any WASH intervention. In measurements with above median weekly total rainfall, we estimated a prevalence ratio of 0.56 (95% CI 0.44-0.72) for any WASH intervention compared to 0.85 (95% CI 0.42-1.70) in measurements with below median total rainfall. (Figure 3, Table A1) The prevalence ratio for the pooled WASH intervention was 0.49 (95% CI 0.40-0.61) during the rainy season compared to 1.06 (95% CI 0.75-1.51) during the dry season (Figure 3, Table A1). The prevalence ratio associated with any WASH intervention was lower following weeks when there was at least one day of heavy rainfall (0.49, 95% CI 0.35-0.68) compared to when there were no days with heavy rainfall (0.86, 95% CI 0.60-1.25) (Figure 3, Table A1).

**Figure 3:**
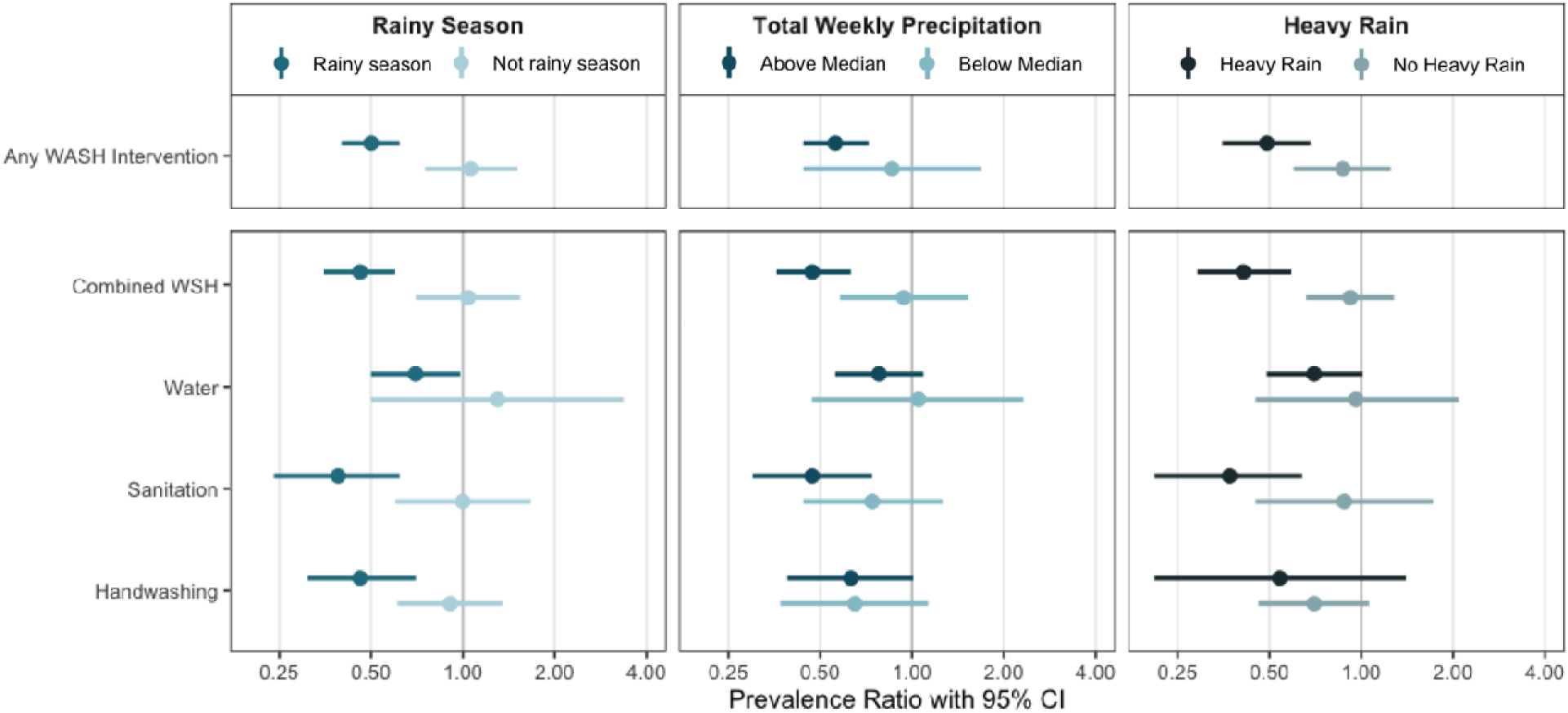
Differences in diarrhea prevalence under varying precipitation conditions, by intervention group. Prevalence ratios for caregiver-reported diarrhea in the intervention vs control groups. Heavy rain describes whether there is at least 1 day with over 80^th^ percentile daily rainfall in the 8-14 day prior to outcome assessment. Total precipitation is also measured for the 8-14 day period prior to outcome assessment, with a median value of 13 mm. All effect estimates have controlled for average weekly temperature.

Next, we assessed effect modification by precipitation for each intervention type (Figure 3). During the rainy season, the sanitation, handwashing, combined WSH interventions reduced diarrhea prevalence by 54% to 61%, while the water intervention reduced it by 30% (Figure 3). For all intervention arms, there was no decrease in diarrhea prevalence in the dry season. Intervention-specific trends were similar for total precipitation and heavy rainfall, with stronger effect modification for heavy rain than for total weekly precipitation. However, for the handwashing intervention, there was no evidence of effect modification by total weekly precipitation or heavy rainfall.

### Temperature

During the study, the weekly average temperature ranged from 18 to 32°C (median = 27°C), the minimum temperature ranged from 17 to 31°C (median = 25°C), and the maximum temperature ranged from 18 to 34°C (median = 28°C). Temperatures reached their peak in May, immediately preceding or at the start of the annual rainy season. (Figure 1C)

We found that as average temperature increased, diarrhea prevalence increased slightly in the intervention arms but increased rapidly in the control arm (Figure 4A). At an average temperature of 20°C (10^th^ percentile), the prevalence ratio for any WASH intervention was 1.76 (95% CI 0.96-3) compared to 0.84 (95% CI 0.47-1.18) at an average temperature of 30°C (90^th^ percentile). We saw similar increases in intervention effectiveness under higher minimum temperatures (10^th^ percentile PR = 1.72, 95% CI 0.96-2.92 at 19°C vs 90^th^ percentile PR = 0.84, 95% CI 0.46-1.16 at 29°C) and maximum temperatures (10^th^ percentile PR = 1.90, 95% CI 1.05-3.30 at 22°C vs 90^th^ percentile PR = 0.90, 95% CI 0.59-1.28 at 32°C). We saw similar trends when comparing above median versus below median measurements and estimated prevalence ratios of 0.59 (95% CI 0.48-0.74) versus 0.91 (95% CI 0.60-1.36) for average temperatures, 0.63 (95% CI 0.50-0.79) vs 0.84 (95% CI 0.40-1.76) for minimum temperatures, and 0.60 (95% CI 0.47-0.75) vs 0.91 (95% CI 0.62-1.33) for maximum temperatures (Figure 4B).

**Figure 4:**
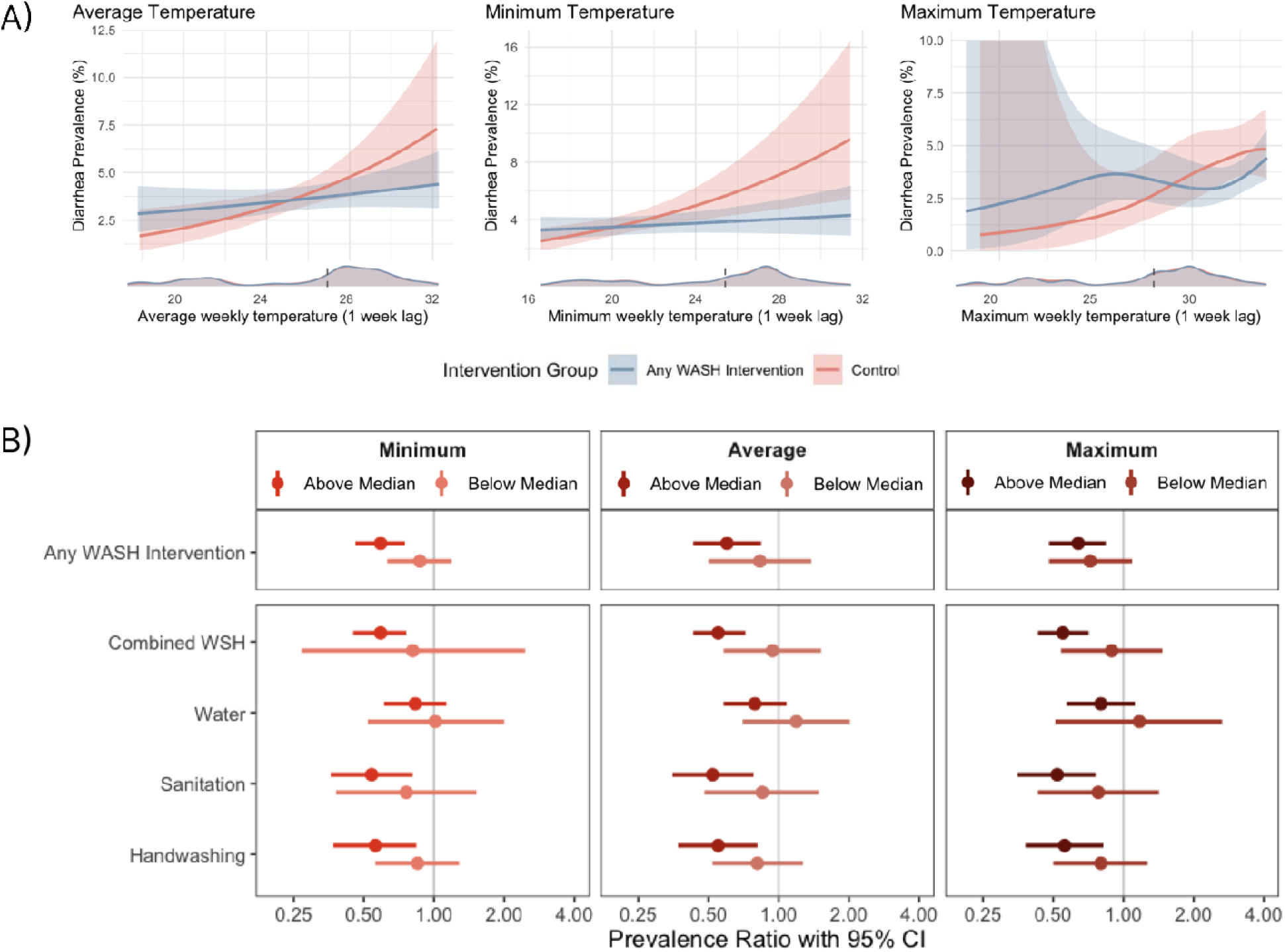
Differences in diarrhea prevalence under varying temperature conditions, by intervention group. (A) Predicted diarrhea prevalence by average, minimum, and maximum temperatures in the 8-14 day period prior to outcome assessment, controlled for total weekly precipitation. Density plots show the distribution of measurements over values of total temperature, with a dashed line marking the median. (B) Prevalence ratios for caregiver-reported diarrhea in the intervention vs control groups. All effect estimates have controlled for total weekly precipitation. The median values were 27°C for average temperature, 25°C for minimum temperature, and 28°C for maximum temperature.

Temperature appeared to have a larger influence on the effectiveness of the sanitation, handwashing, and combined WSH interventions compared to the water intervention. (Figure 4B, Table A2) During periods in which there were above median weekly average temperatures, we estimated between a 41% to 46% reduction in diarrhea under the sanitation, handwashing, and combined WSH interventions compared to a 17% reduction under the water intervention. We found no difference in effectiveness of the water, sanitation, or handwashing interventions by minimum of maximum temperatures, though the below median temperature effects were null and the above median effects were not. However, our estimates have low precision.

### Other analyses

We did not observe any significant differences in intervention effect estimates by climate or environment on caregiver-reported bruising, a negative-control outcome. (Appendix 2)

We conducted sensitivity analyses under different lag periods for precipitation and temperature. Estimates of intervention effect across different precipitation (Figure A1) and temperature (Figure A2) conditions were consistent across all lag periods we assessed.

## Discussion

WASH interventions in rural Bangladesh more effectively prevented child diarrhea under high temperatures and precipitation. We found that receipt of any WASH intervention reduced diarrhea prevalence by 51% following heavy rainfall and 41% after above-median temperatures compared to the overall 34% reduction observed in the original trial. We observed no effect of interventions on diarrhea in the dry season or following below-median temperature or precipitation. Effect modification varied by intervention type; precipitation had the strongest influence on the effectiveness of the sanitation and combined WASH interventions while temperature had the strongest influence on the sanitation, handwashing, and combined WASH interventions.

Prior studies have found that diarrhea prevalence in low- and middle-income countries follows a concentration-dilution pattern in which heavy rainfall following dry periods is associated with increased diarrhea risk because pathogens that become concentrated during dry periods are flushed into the environment.^23^ One study also found that drinking water treatments, but not sanitation or hygiene interventions, reduced spikes in diarrhea due to heavy rain following dry periods.^16^ However, during the WASH Benefits trial, there were very few heavy rainfall events preceded by dry periods, and diarrhea prevalence in the control arm was higher following heavy rainfall events, regardless of preceding conditions. Compared to other settings, the annual monsoon in Bangladesh is characterized by higher precipitation intensity and higher total precipitation that is heavily concentrated during the season. Differences in hydrogeology between regions could also impact pathogen transport after rainfall. A study conducted in Indore, India and Kolkata, India found that wells near latrines surrounded by alluvial formations (consisting of loose silt, clay, and sand) had significantly lower fecal coliform and nitrate concentrations compared to wells near latrines with fractured rocks.^24^ In Bangladesh, alluvial sediments are common in flood plains and may have provided a protective barrier that prevented pathogens from being flushed to the surface by heavy rainfall.^25, 26^ Additionally, an analysis of environmental contamination in the WASH Benefits trial found that *E. coli* concentrations in food, stored water, and ponds were elevated following heavy rainfall compared to other periods, but that there was no impact on groundwater quality. (Ercumen A, North Carolina State University, personal communication) These findings suggest that the concentration-dilution pattern does not hold in this setting and increases in diarrhea after heavy rainfall are likely attributed to the contamination of stored water and food supplies.

Interventions were generally more effective under higher temperatures, which might be attributed to increased enteric pathogen transmission in warm conditions. Generally, high temperature in storage containers have been linked to the growth of enteropathogens in food and drinking water supplies.^27, 28^ Ambient temperature may also influence the survival, distribution, and virulence of enteric pathogens in surface water, soil, food, or on surfaces, but few studies have investigated these relationships empirically. An analysis of environmental contamination in the WASH Benefits trial provides evidence that both ambient temperature and higher temperature in storage containers are associated with higher pathogen concentrations: during or following extreme heat, *E. coli* concentrations in food, stored water, ponds, and soil were elevated 1.25-2 fold compared to cooler periods. (Ercumen A, North Carolina State University, personal communication) A prior meta-analysis found that increased temperatures were strongly associated with increased all-cause and bacterial diarrhea, but not viral diarrhea.^6^ Bacterial pathogens are a common cause of diarrhea in children <2 years in Bangladesh,^29^ and it is possible that associations between diarrhea and temperature in this study were due to increased transmission of bacterial but not viral enteropathogens. However, we did not investigate diarrhea etiology in this study.

Bangladesh is projected to experience rapidly increasing temperatures and precipitation under climate change, which may increase the burden of childhood diarrhea in rural communities. Our findings suggest that WASH interventions will be particularly impactful for preventing diarrhea under more extreme weather conditions and may be resistant to damages during typical monsoon season. As communities brace for the impacts of climate change, investment into WASH interventions may increase the resilience of vulnerable populations against diarrhea. We found that the sanitation intervention more effectively mitigated increases in diarrhea prevalence following precipitation and higher temperatures, and the water intervention had the smallest preventive effect on diarrhea in these conditions. This may be because the sanitation intervention interrupts multiple upstream pathways of enteric pathogen transmission through flies, water, or soil, while the handwashing and water interventions interrupt fewer pathways that are more downstream. To mitigate the impacts of climate change on diarrhea, interventions that address multiple pathways of enteric pathogen transmission may be necessary.

Our study is subject to several limitations. We measured caregiver-reported diarrhea, which is susceptible to courtesy bias. However, our negative control analysis using an alternative caregiver-reported outcome suggested that there was no evidence of misclassification. Second, we were not able to investigate the influence of flooding due to a lack of available data, and we could not investigate the interaction between heavy rain preceded by dry periods due to data sparsity. Third, the hottest temperatures during the study coincided with the end of the dry season, so the impact of temperature was difficult to disentangle from that of season. Finally, because higher temperature and higher precipitation mostly coincided during the study period, it was difficult to fully isolate the influence of each on WASH intervention effectiveness.

## Conclusion

Here, we rigorously assessed the influence of temperature and precipitation on WASH intervention effectiveness using data from randomized trial with high adherence and high-resolution weather data. Low-cost, household level WASH interventions more effectively reduced diarrhea prevalence following periods of higher temperatures, higher precipitation, and heavy rainfall. Effect modification varied by intervention type, and we observed the largest differences in diarrhea reductions following heavy rainfall under the sanitation intervention. In regions with similar climates, WASH interventions may increase community resilience against extreme weather under climate change by preventing environmentally mediated enteric infections.

## Acknowledgements

This study was supported by the Gates Foundation (grant number OPPGD759 to the University of California, Berkeley). The original trial was implemented by icddr,b, and we greatly appreciate the contributions of the study participants and field workers who delivered the interventions and led data collection. Research reported in this publication was supported in part by the National Institute of Allergy And Infectious Diseases of the National Institutes of Health under Award Numbers K01AI141616 (PI: Benjamin-Chung) and R01AI166671 (PI: Arnold), the National Heart, Lung, And Blood Institute of the National Institutes of Health under award number T32HL151323 (Nguyen), the National Institute of General Medical Sciences of the National Institutes of Health under award number T32GM144273 (Athni), and a Stanford University School of Medicine Dean’s Postdoctoral Fellowship (Grembi). The content is solely the responsibility of the authors and does not necessarily represent the official views of the National Institutes of Health. Jade Benjamin-Chung is a Chan Zuckerberg Biohub Investigator. We also acknowledge the Stanford Research Computing Center for computational resources at the Sherlock high-performance cluster.

## Data Sharing

Individual participant data and metadata for this study will be made available at the time of publication and posted here: https://osf.io/yt67k/ The pre-analysis plan and ancillary results are also available at the same URL. To protect participant privacy, household geocoordinates are not included in the public dataset.

## Notes

### Competing Interest Statement

The authors have declared no competing interest.

### Clinical Trial

NCC01590095

### Funding Statement

This study was supported by the Gates Foundation (grant number OPPGD759 to the University of California, Berkeley). Research reported in this publication was supported in part by the National Institute of Allergy And Infectious Diseases of the National Institutes of Health under Award Numbers K01AI141616 (PI: Benjamin-Chung) and R01AI166671 (PI: Arnold), the National Heart, Lung, And Blood Institute of the National Institutes of Health under award number T32HL151323 (Nguyen), the National Institute of General Medical Sciences of the National Institutes of Health under award number T32GM144273 (Athni), and a Stanford University School of Medicine Dean's Postdoctoral Fellowship (Grembi). The content is solely the responsibility of the authors and does not necessarily represent the official views of the National Institutes of Health. Jade Benjamin-Chung is a Chan Zuckerberg Biohub Investigator.

### Author Declarations

The original trial protocol was approved by the Ethical Review Committee at The International Centre for Diarrhoeal Disease Research, Bangladesh (PR-11063), the Committee for the Protection of Human Subjects at the University of California, Berkeley (2011-09-3652), and the institutional review board at Stanford University (25863).

### Summary of Updates

Removed climate change projections, added arm-specific analyses, focused on specific lag periods

