## Supplementary Materials for "Influence of temperature and precipitation on the effectiveness of water, sanitation, and handwashing interventions against childhood diarrheal disease in rural Bangladesh: a re-analysis of a randomized control trial"

**Appendix 1: Deviations from the pre-analysis plan**

Prior to this study, we published a pre-analysis plan at <https://osf.io/yt67k/>. Any deviations that we made to this plan are listed below.

1. We pre-specified an analysis that included an interaction between weekly precipitation measurements and whether there was rain in the prior 60 days. This had been done in prior studies to differentiate rainfall that followed a dry versus wet period. We were not able to conduct this analysis because high levels of rainfall almost never followed a dry period in this study region.
2. We pre-specified the use of the NASA MOD11A1 temperature dataset. However, we found that this data had a large proportion of missing values that were concentrated during the rainy season, when cloud coverage may have impacted quality of measurements. Instead, we used the FLDAS dataset that was more complete but had lower spatial granularity.
3. We pre-specified the measurement of average, minimum, and maximum temperatures in the 1-day, 7-day, 30-day, and 90-day periods prior to outcome assessment. We decided to use lagged weekly variables instead to be consistent with our precipitation measurements. Additionally, we constructed a binary “extreme temperature” variable to capture the potential effects of heat waves to reflect what has been studied in existing literature.
4. We did not pre-specify the calculation of prevalence ratios at the 10^th^ and 90^th^ percentile values of the continuous risk factor variables.
5. We ran analyses for water flow accumulation, the enhanced vegetative index, land use, and population density per our pre-analysis plan. Overall, we did not observe associations with study outcomes. We excluded them from this manuscript. Results are available here: https://osf.io/yt67k/

**Appendix 2: Negative Control Analysis**

We conducted a negative control analysis by estimating effect modification of WASH interventions on caregiver-reported bruising by all categorical measures of temperature and precipitation under a 1-week lag.

There is no evidence of residual confounding, and we can reasonably assume that the caregiver reported measured of diarrhea is valid.

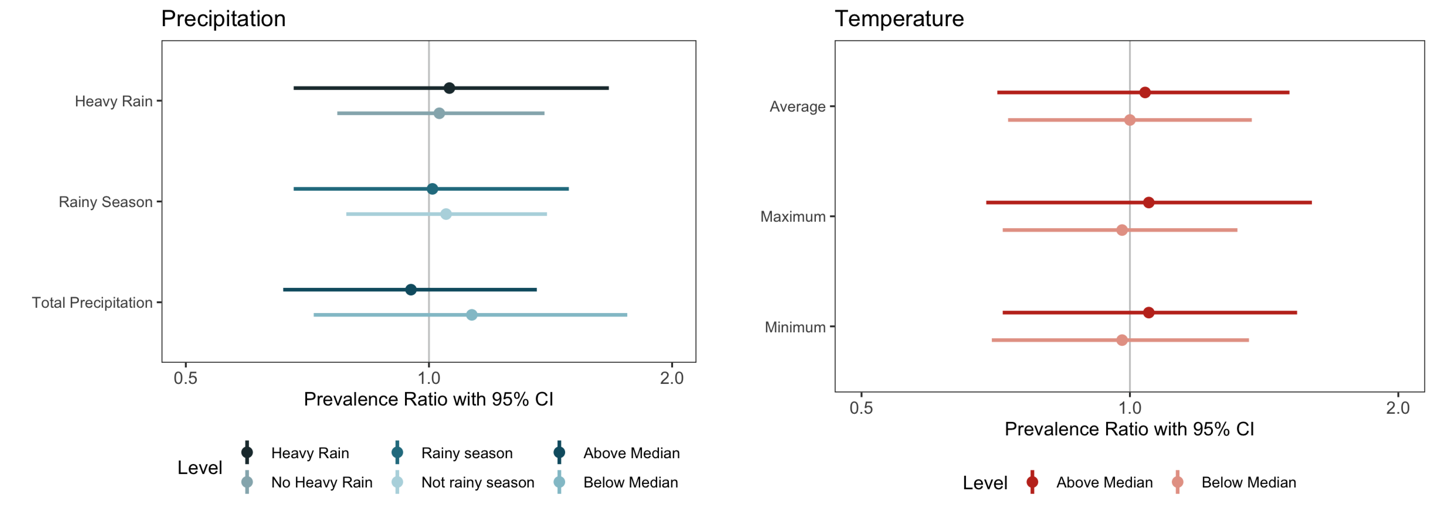

**Figure A1: Sensitivity Analysis - WASH intervention effectiveness by precipitation under varying lag periods**

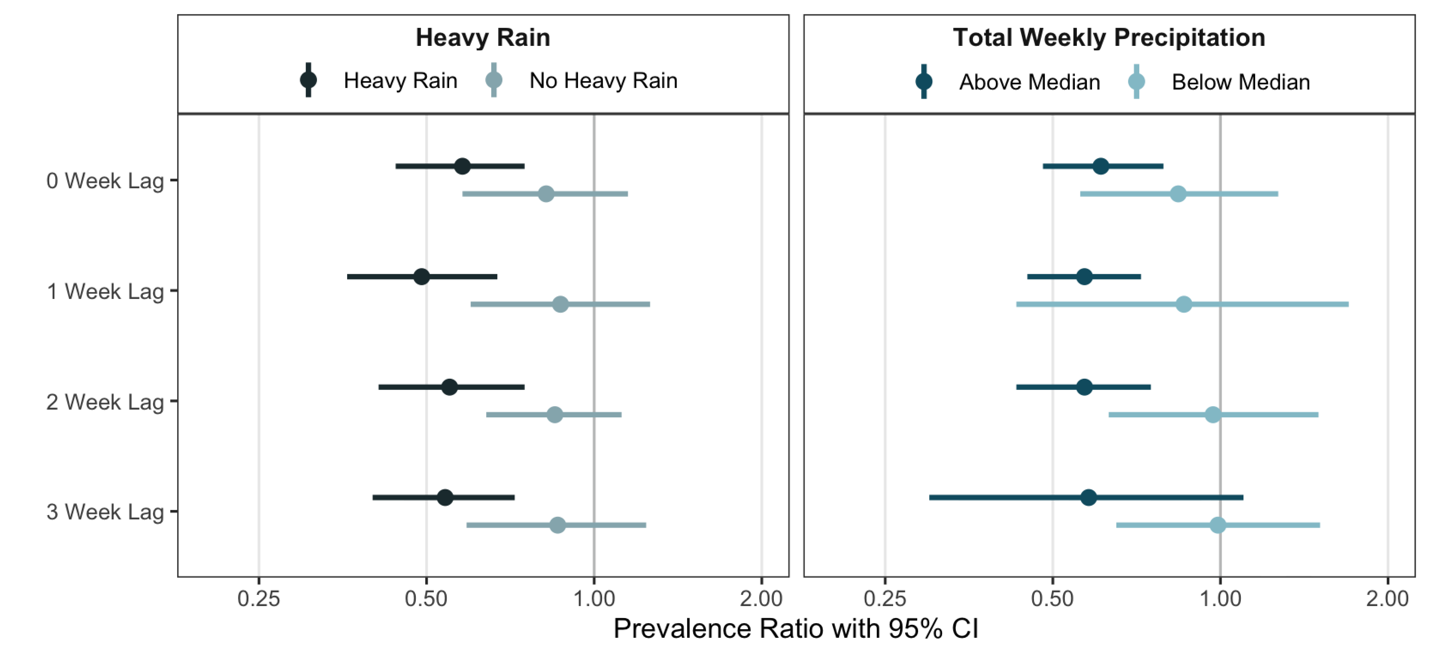

**Figure A2: Sensitivity Analysis - WASH intervention effectiveness by temperature under varying lag periods**

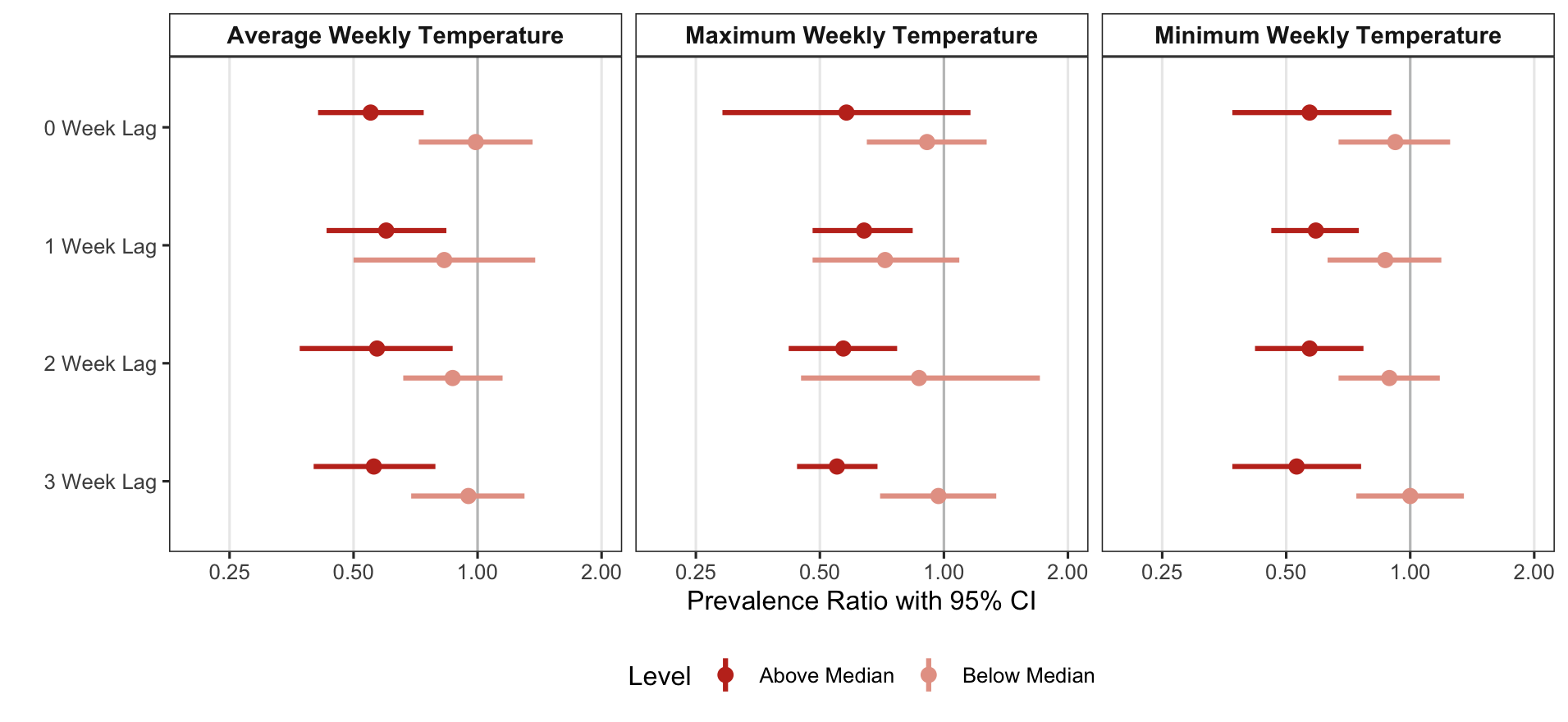

**Table A1: Estimated Prevalence Ratios Under Different Precipitation Conditions**

| **Variable** | **Level** | **N** | **Risk in Intervention Arm** | **Risk in Control Arm** | **Risk Ratio (95% CI)** | **Risk Difference (95% CI)** |
| --- | --- | --- | --- | --- | --- | --- |
| ***Pooled Intervention*** | | | | | | |
| Rainy Season | Rainy season | 6,350 | 0.04 (0.03, 0.04) | 0.08 (0.06, 0.09) | 0.49 (0.40, 0.62) | -0.04 (-0.05, -0.02) |
|  | Not rainy season | 6,090 | 0.04 (0.03, 0.04) | 0.04 (0.03, 0.04) | 1.06 (0.75, 1.51) | 0.00 (-0.01, 0.02) |
| Heavy Rain | Heavy Rain | 5,473 | 0.03 (0.03, 0.04) | 0.07 (0.05, 0.08) | 0.49 (0.36, 0.67) | -0.03 (-0.05, -0.02) |
|  | No Heavy Rain | 6,967 | 0.04 (0.04, 0.05) | 0.04 (0.04, 0.05) | 0.87 (0.60, 1.26) | -0.01 (-0.02, 0.01) |
| Total Weekly Precipitation | Above Median | 7,192 | 0.04 (0.03, 0.04) | 0.06 (0.05, 0.08) | 0.57 (0.45, 0.72) | -0.03 (-0.04, -0.01) |
|  | Below Median | 5,248 | 0.04 (0.03, 0.05) | 0.04 (0.03, 0.05) | 0.86 (0.43, 1.70) | -0.01 (-0.04, 0.02) |
| ***Combined WSH*** | | | | | | |
| Rainy Season | Rainy season | 3,675 | 0.03 (0.03, 0.04) | 0.08 (0.06, 0.09) | 0.46 (0.35, 0.60) | -0.04 (-0.06, -0.03) |
|  | Not rainy season | 3,481 | 0.04 (0.03, 0.05) | 0.04 (0.03, 0.05) | 1.04 (0.70, 1.54) | 0.00 (-0.01, 0.02) |
| Heavy Rain | Heavy Rain | 3,139 | 0.03 (0.02, 0.04) | 0.07 (0.05, 0.08) | 0.41 (0.29, 0.59) | -0.04 (-0.05, -0.02) |
|  | No Heavy Rain | 4,017 | 0.04 (0.03, 0.05) | 0.04 (0.03, 0.05) | 0.92 (0.66, 1.28) | 0.00 (-0.02, 0.01) |
| Total Weekly Precipitation | Above Median | 4,161 | 0.03 (0.02, 0.04) | 0.06 (0.05, 0.08) | 0.47 (0.35, 0.62) | -0.03 (-0.05, -0.02) |
|  | Below Median | 2,995 | 0.04 (0.03, 0.06) | 0.04 (0.03, 0.06) | 0.95 (0.59, 1.53) | 0.00 (-0.02, 0.02) |
| ***Water*** | | | | | | |
| Rainy Season | Rainy season | 2,687 | 0.05 (0.04, 0.07) | 0.08 (0.06, 0.09) | 0.70 (0.51, 0.98) | -0.02 (-0.04, 0.00) |
|  | Not rainy season | 2,569 | 0.05 (0.03, 0.06) | 0.05 (0.03, 0.06) | 1.30 (0.50, 3.37) | 0.01 (-0.03, 0.05) |
| Heavy Rain | Heavy Rain | 2,357 | 0.05 (0.03, 0.07) | 0.07 (0.05, 0.08) | 0.70 (0.49, 1.00) | -0.02 (-0.04, 0.00) |
|  | No Heavy Rain | 2,899 | 0.05 (0.04, 0.06) | 0.05 (0.04, 0.06) | 0.98 (0.47, 2.06) | 0.00 (-0.04, 0.03) |
| Total Weekly Precipitation | Above Median | 3,090 | 0.05 (0.04, 0.06) | 0.06 (0.05, 0.08) | 0.78 (0.56, 1.09) | -0.01 (-0.03, 0.00) |
|  | Below Median | 2,166 | 0.05 (0.03, 0.07) | 0.05 (0.03, 0.07) | 1.05 (0.46, 2.37) | 0.00 (-0.04, 0.04) |
| ***Sanitation*** | | | | | | |
| Rainy Season | Rainy season | 2,649 | 0.03 (0.02, 0.04) | 0.08 (0.06, 0.09) | 0.39 (0.24, 0.62) | -0.05 (-0.07, -0.03) |
|  | Not rainy season | 2,547 | 0.04 (0.02, 0.05) | 0.04 (0.02, 0.05) | 1.00 (0.60, 1.68) | 0.00 (-0.02, 0.02) |
| Heavy Rain | Heavy Rain | 2,295 | 0.03 (0.01, 0.04) | 0.07 (0.05, 0.08) | 0.37 (0.21, 0.64) | -0.04 (-0.07, -0.02) |
|  | No Heavy Rain | 2,901 | 0.04 (0.03, 0.05) | 0.04 (0.03, 0.05) | 0.88 (0.45, 1.72) | -0.01 (-0.04, 0.02) |
| Total Weekly Precipitation | Above Median | 2,981 | 0.03 (0.02, 0.04) | 0.06 (0.05, 0.08) | 0.47 (0.30, 0.74) | -0.03 (-0.05, -0.01) |
|  | Below Median | 2,215 | 0.04 (0.02, 0.06) | 0.04 (0.02, 0.06) | 0.75 (0.44, 1.27) | -0.01 (-0.03, 0.01) |
| ***Handwashing*** | | | | | | |
| Rainy Season | Rainy season | 2,655 | 0.04 (0.02, 0.05) | 0.08 (0.06, 0.09) | 0.46 (0.30, 0.70) | -0.04 (-0.06, -0.02) |
|  | Not rainy season | 2,575 | 0.03 (0.02, 0.04) | 0.03 (0.02, 0.04) | 0.91 (0.61, 1.35) | 0.00 (-0.02, 0.01) |
| Heavy Rain | Heavy Rain | 2,275 | 0.04 (0.02, 0.05) | 0.07 (0.05, 0.08) | 0.54 (0.21, 1.38) | -0.03 (-0.07, 0.00) |
|  | No Heavy Rain | 2,955 | 0.03 (0.02, 0.04) | 0.03 (0.02, 0.04) | 0.69 (0.45, 1.06) | -0.01 (-0.03, 0.00) |
| Total Weekly Precipitation | Above Median | 2,987 | 0.04 (0.03, 0.05) | 0.06 (0.05, 0.08) | 0.63 (0.39, 1.02) | -0.02 (-0.04, 0.00) |
|  | Below Median | 2,243 | 0.03 (0.02, 0.04) | 0.03 (0.02, 0.04) | 0.65 (0.37, 1.12) | -0.02 (-0.04, 0.01) |

**Table A2: Estimated Prevalence Ratios Under Different Temperature Conditions**

| **Variable** | **Level** | **N** | **Risk in Intervention Arm** | **Risk in Control Arm** | **Risk Ratio (95% CI)** | **Risk Difference (95% CI)** |
| --- | --- | --- | --- | --- | --- | --- |
| ***Pooled Intervention*** | | | | | | |
| Average Weekly Temperature | Above Median | 7,329 | 0.04 (0.03, 0.05) | 0.07 (0.05, 0.08) | 0.60 (0.43, 0.84) | -0.03 (-0.04, -0.01) |
|  | Below Median | 5,111 | 0.04 (0.03, 0.04) | 0.04 (0.03, 0.04) | 0.83 (0.50, 1.38) | -0.01 (-0.03, 0.02) |
| Minimum Weekly Temperature | Above Median | 7,358 | 0.04 (0.03, 0.04) | 0.07 (0.05, 0.08) | 0.59 (0.46, 0.75) | -0.03 (-0.04, -0.01) |
|  | Below Median | 5,082 | 0.04 (0.03, 0.04) | 0.04 (0.03, 0.04) | 0.87 (0.63, 1.19) | -0.01 (-0.02, 0.01) |
| Maximum Weekly Temperature | Above Median | 6,700 | 0.04 (0.04, 0.05) | 0.07 (0.05, 0.08) | 0.64 (0.48, 0.84) | -0.02 (-0.04, -0.01) |
|  | Below Median | 5,740 | 0.03 (0.03, 0.04) | 0.03 (0.03, 0.04) | 0.72 (0.48, 1.09) | -0.01 (-0.03, 0.01) |
| ***Combined WSH*** | | | | | | |
| Average Weekly Temperature | Above Median | 4,542 | 0.04 (0.03, 0.05) | 0.07 (0.05, 0.08) | 0.55 (0.43, 0.72) | -0.03 (-0.04, -0.02) |
|  | Below Median | 2,576 | 0.03 (0.02, 0.04) | 0.03 (0.02, 0.04) | 0.94 (0.58, 1.52) | 0.00 (-0.02, 0.01) |
| Minimum Weekly Temperature | Above Median | 4,603 | 0.04 (0.03, 0.05) | 0.07 (0.05, 0.08) | 0.59 (0.45, 0.76) | -0.03 (-0.04, -0.01) |
|  | Below Median | 2,515 | 0.03 (0.02, 0.04) | 0.03 (0.02, 0.04) | 0.81 (0.27, 2.46) | -0.01 (-0.05, 0.03) |
| Maximum Weekly Temperature | Above Median | 4,441 | 0.04 (0.03, 0.05) | 0.07 (0.06, 0.08) | 0.55 (0.43, 0.71) | -0.03 (-0.05, -0.02) |
|  | Below Median | 2,677 | 0.03 (0.02, 0.04) | 0.03 (0.02, 0.04) | 0.89 (0.54, 1.47) | 0.00 (-0.02, 0.01) |
| ***Water*** | | | | | | |
| Average Weekly Temperature | Above Median | 3,276 | 0.05 (0.04, 0.07) | 0.07 (0.05, 0.08) | 0.79 (0.58, 1.08) | -0.01 (-0.03, 0.00) |
|  | Below Median | 1,970 | 0.04 (0.03, 0.06) | 0.04 (0.03, 0.06) | 1.19 (0.70, 2.02) | 0.01 (-0.01, 0.03) |
| Minimum Weekly Temperature | Above Median | 3,367 | 0.05 (0.04, 0.07) | 0.07 (0.05, 0.08) | 0.83 (0.61, 1.13) | -0.01 (-0.03, 0.01) |
|  | Below Median | 1,879 | 0.04 (0.03, 0.06) | 0.04 (0.03, 0.06) | 1.01 (0.52, 2.00) | 0.00 (-0.03, 0.03) |
| Maximum Weekly Temperature | Above Median | 3,213 | 0.06 (0.04, 0.07) | 0.07 (0.06, 0.08) | 0.80 (0.57, 1.12) | -0.01 (-0.03, 0.01) |
|  | Below Median | 2,033 | 0.04 (0.02, 0.06) | 0.04 (0.02, 0.06) | 1.17 (0.51, 2.65) | 0.01 (-0.03, 0.04) |
| ***Sanitation*** | | | | | | |
| Average Weekly Temperature | Above Median | 3,209 | 0.04 (0.02, 0.05) | 0.07 (0.05, 0.08) | 0.52 (0.35, 0.78) | -0.03 (-0.05, -0.01) |
|  | Below Median | 1,977 | 0.03 (0.02, 0.04) | 0.03 (0.02, 0.04) | 0.85 (0.48, 1.49) | -0.01 (-0.02, 0.01) |
| Minimum Weekly Temperature | Above Median | 3,296 | 0.04 (0.02, 0.05) | 0.07 (0.05, 0.08) | 0.54 (0.36, 0.81) | -0.03 (-0.05, -0.01) |
|  | Below Median | 1,890 | 0.03 (0.02, 0.04) | 0.03 (0.02, 0.04) | 0.76 (0.38, 1.52) | -0.01 (-0.03, 0.01) |
| Maximum Weekly Temperature | Above Median | 3,171 | 0.04 (0.02, 0.05) | 0.07 (0.06, 0.08) | 0.52 (0.35, 0.76) | -0.03 (-0.05, -0.02) |
|  | Below Median | 2,015 | 0.03 (0.01, 0.04) | 0.03 (0.01, 0.04) | 0.78 (0.43, 1.42) | -0.01 (-0.03, 0.01) |
| ***Handwashing*** | | | | | | |
| Average Weekly Temperature | Above Median | 3,227 | 0.04 (0.03, 0.05) | 0.07 (0.05, 0.08) | 0.55 (0.37, 0.81) | -0.03 (-0.05, -0.01) |
|  | Below Median | 1,993 | 0.03 (0.02, 0.04) | 0.03 (0.02, 0.04) | 0.81 (0.52, 1.27) | -0.01 (-0.02, 0.01) |
| Minimum Weekly Temperature | Above Median | 3,315 | 0.04 (0.03, 0.05) | 0.07 (0.05, 0.08) | 0.56 (0.37, 0.84) | -0.03 (-0.05, -0.01) |
|  | Below Median | 1,905 | 0.03 (0.02, 0.04) | 0.03 (0.02, 0.04) | 0.85 (0.56, 1.28) | -0.01 (-0.02, 0.01) |
| Maximum Weekly Temperature | Above Median | 3,161 | 0.04 (0.03, 0.05) | 0.07 (0.06, 0.08) | 0.56 (0.38, 0.82) | -0.03 (-0.05, -0.01) |
|  | Below Median | 2,059 | 0.03 (0.02, 0.04) | 0.03 (0.02, 0.04) | 0.80 (0.50, 1.27) | -0.01 (-0.02, 0.01) |
